# Chronic absenteeism in Canadian kindergarten classes, pre- and post-COVID-19, and its association with concurrent developmental vulnerability

**DOI:** 10.64898/2026.03.04.26347661

**Authors:** Caroline Reid-Westoby, Eric Duku, Ashley Gaskin, Magdalena Janus

**Affiliations:** Offord Centre for Child Studies, Department of Psychiatry and Behavioural Neurosciences, McMaster University, Hamilton, Ontario, Canada; Department of Pediatrics, Faculty of Medicine, University of British Columbia, Vancouver, British Columbia, Canada

**Keywords:** school readiness, chronic absenteeism, COVID-19, kindergarten, developmental health, Canada

## Abstract

Students who frequently miss school are at greater risk for academic difficulty. High levels of absenteeism as early as kindergarten have been associated with long-term consequences, such as low reading proficiency in Grade 3 and low academic achievement in Grade 5, both of which have been associated with lower rates of high school graduation and enrollment in post-secondary education. The prevalence of school absenteeism has increased significantly since the COVID-19 pandemic and there have been sustained shifts in student attendance rates from kindergarten to Grade 12 since 2020. The goals of this population-level, repeated cross-sectional cohort study were to compare rates of chronic absenteeism, defined as being absent from school at least 10% of the time, in kindergarten in Canada before and after the onset of the COVID-19 pandemic, and examine the association between children’s chronic absenteeism and their concurrent developmental vulnerability. A total of 513,159 kindergarten children participated in the study, with 284,712 (55.5%) being in the pre-COVID-19 cohort (2017–2020) and 228,447 (44.5%) in the post-COVID-19 cohort (2020–2023). Across Canada, rates of chronic absenteeism increased from pre- to post-COVID-19, from 17.7% to 41.3%, with differences by jurisdiction. The greatest increase was seen in Ontario, while the smallest increase was seen in British Columbia. Children attending kindergarten in the post-COVID-19 cohort were three times more likely to be chronically absent compared to their peers attending kindergarten before the onset of the pandemic. Despite this, chronic absenteeism in the post-COVID-19 period was associated with reduced odds of overall developmental vulnerability, a pattern that is likely attributable to shifts in the composition of chronically absent children. In the post-COVID-19 cohort, a greater percentage of children who were chronically absent resided in higher SES neighbourhoods compared to their chronically absent peers attending school before the onset of the pandemic. While increasing rates of school absenteeism should not be ignored, our results suggest that chronic absenteeism following COVID-19 might be more nuanced than before. The jurisdictional differences in rates of chronic absenteeism observed in this study could be due to the various public health measures put in place by the various provincial and territorial governments. It is also possible that the children from higher SES neighbourhoods missed more school after the onset of the COVID-19 pandemic because their parents had the capability to work from home, making it easier to keep their child(ren) home from school. The decreased association between chronic absenteeism and developmental vulnerability post-COVID-19 may reflect improved access to online resources, which enables students to stay on track academically from home. Gaining a better understanding of the reasons behind missing school and the relation between absenteeism and academic achievement at various developmental stages is crucial to support successful learning trajectories.

## Introduction

Early school experiences play an important role in a child’s life, as they help foster engagement with the educational process. School engagement has been associated with future academic and social success (1). A key aspect of being engaged is regular school attendance. Research suggests that one benefit of attending school is a function of exposure: greater exposure (i.e. attendance) leads to increased educational gains, whereas less exposure (i.e. frequent absences) can hinder learning and result in fewer gains (2). Students who are more frequently absent are at greater risk for school difficulty (3–7). High levels of absenteeism as early as kindergarten are associated with long-term consequences such as low reading proficiency in Grade 3 (8) and low academic achievement in Grade 5 (9,10), both of which have been associated with lower rates of high school graduation and enrollment in post-secondary education.

Some experts maintain that children’s early life experiences are particularly crucial for their long-term school performance (11) as the skills and knowledge learned during these early years lay the groundwork for what they will learn in the future (1). Consequently, if children are not attending school during this formative time, they may be less likely to succeed later on. Only a small number of studies has looked at the associations between school absences and other dimensions of development, other than academic attainment, such as socioemotional skills. Gottfried et al. (5), for example, found that kindergartners who were more frequently absent exhibited less optimal social skills and greater internalizing problems at the end of the school year. Research has shown that children who frequently miss school are at increased risk of poor academic achievement and socioemotional problems, risky health behaviours, discontinuing education, not attending post-secondary education, as well as lower income, frequent work absences, and poorer health as adults (5,12,13).

School absenteeism starts early and may persist over time (14,15). Rates of school absenteeism appear to be highest in kindergarten (16,17). Approximately 10% of children enrolled in kindergarten in the United States are chronically absent, defined as missing school at least 10% of the time (10). A national survey in the United States found that, on average, 14% of students are chronically absent, with rates varying by jurisdiction from less than 5% to more than 50% (18). Frequent school absences are often driven by many factors, with illness being the most commonly reported reason for missing school. The prevalence of absenteeism is higher in students with disabilities or low academic achievement (16). Nevertheless, not all children who are repeatedly absent have a chronic disease or are ill. A number of studies have shown that child, family, community, and school characteristics all contribute to school absenteeism (19–24).

Another factor associated with school absenteeism is the COVID-19 pandemic. The prevalence of school absenteeism has increased significantly since the COVID-19 pandemic and there have been sustained shifts in student attendance rates from kindergarten to Grade 12 since 2020. The percentage of public-school students in the United States who were chronically absent almost doubled between the 2018–19 and 2021–22 school years, from 15% to 28% (25). In a study in North Carolina, the percentage of students who were chronically absent at least once over the 3-year period increased from 17% pre-pandemic to 38% post-pandemic, while the percentage who were chronically absent in all 3 years quadrupled from 2.4% to 9.6% (26). Even after public health measures subsided, rates of chronic absenteeism have remained high (27). While there is clear evidence that school absenteeism rates have increased since the onset of the COVID-19 pandemic, to our knowledge, no research has examined the association between school absenteeism and children’s concurrent development health, pre- and post-COVID-19. More broadly, research on chronic absenteeism in Canada remains limited.

### Current study

The goal of the current study was to examine the chronic absenteeism rates among Canadian kindergartners before and after the onset of the COVID-19 pandemic and the associations with their school readiness.

This was done by conducting analyses to answer the following research questions:

1) Did rates of chronic absenteeism change in Canadian kindergarten students from pre- to post-COVID-19 onset?
2) Has the demographic composition of chronically absent children changed between the pre- and post-COVID-19 cohorts of Canadian kindergarten children?
3) Has the association between chronic absenteeism and children’s developmental vulnerability changed between the pre- and post-COVID-19 cohorts?

Examining this association in kindergarten children pre- and post-COVID-19 is important, as development at that age is associated with future academic and socioemotional outcomes (8,28,29). Previous research has established that the numbers of days absent from school have increased since the onset of COVID-19 (30), however, it is not known is whether the association between school absenteeism and children’s development and school readiness has changed. In fact, very little research on chronic absenteeism has been conducted in Canada, in any age group, potentially due to the difficulty in acquiring information on school absenteeism (31).

## Methods

### Study Population and Design

This was a population-level, repeated cross-sectional cohort study which consisted of two population-level cohorts of children, one who attended kindergarten before the onset of COVID-19 and another one attending kindergarten post-COVID-19. The study population included for children attending publicly-funded schools from 8 of Canada’s 13 provinces and territories.^1^ The kindergarten teachers completed the Early Development Instrument (EDI) for every child in their classroom between 2017 and 2020 (pre-COVID-19) and 2020 and 2023 (post-COVID-19). Data accessed for this study were de-identified during data collection. Inclusion criteria were being enrolled in kindergarten, being in their current classroom for at least one month, having an EDI questionnaire with no more than 25% of items missing, and not missing data for any of the variables of interest.

### Measures

This study utilized data from the EDI, a 103-item, teacher-completed measure of children’s abilities to meet age-appropriate abilities and behaviours (33). As a measure of child’s developmental health, the EDI questions focus on developmental skills easily observable in the kindergarten classroom crucial for child’s successful transition to primary school. Thus, the EDI outcomes are often conceptualized as school readiness (34).

#### COVID-19 onset

Our study population was divided into two groups: those who attended school before the onset of the COVID-19 pandemic (2017–2020) and those who attended school after the onset of the COVID-19 pandemic (2020–2023). Table 1 lists the provinces and territories included in the current study. A COVID-19 onset variable was created based on when the EDI data was collected in each jurisdiction (pre-COVID-19= 1, post-COVID-19= 2). It should be noted that EDI data collected in 2020 in British Columbia were considered part of the post-COVID-19 pandemic as it was part of their 8th wave of data collection, which included the years 2020, 2021, and 2022. Data collected in Nova Scotia and Northwest Territories in 2020 occurred just before COVID-19 was considered a global pandemic and was therefore included in the pre-COVID-19 cohort.

**Table 1.**
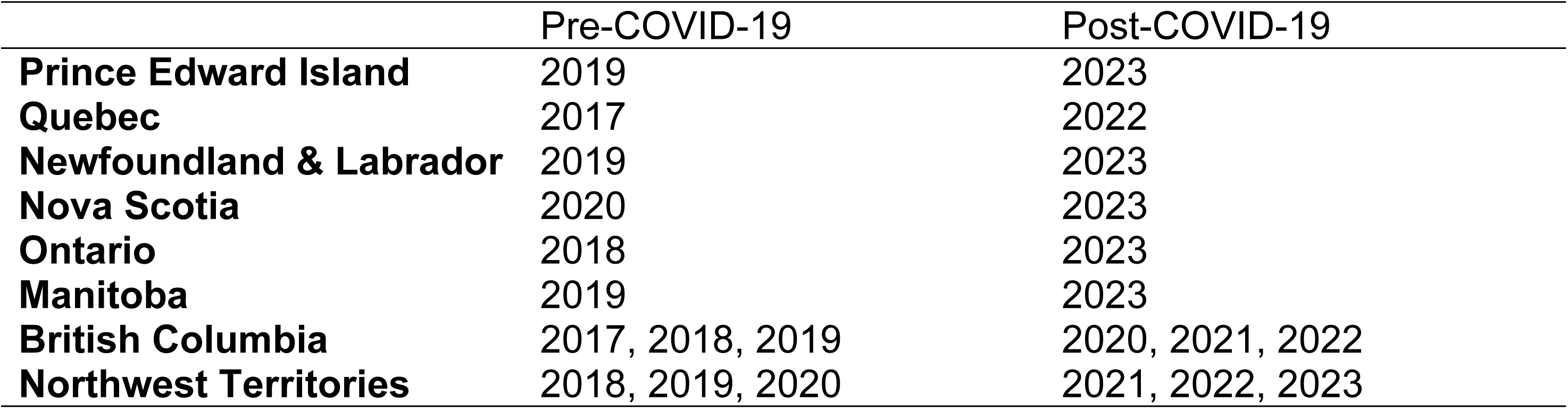
Years of EDI data collections by province and territory, before and after the onset of the COVID-19 pandemic Province/Territory.

|  | Pre-COVID-19 | Post-COVID-19 |
| --- | --- | --- |
| **Prince Edward Island** | 2019 | 2023 |
| **Quebec** | 2017 | 2022 |
| **Newfoundland & Labrador** | 2019 | 2023 |
| **Nova Scotia** | 2020 | 2023 |
| **Ontario** | 2018 | 2023 |
| **Manitoba** | 2019 | 2023 |
| **British Columbia** | 2017, 2018, 2019 | 2020, 2021, 2022 |
| **Northwest Territories** | 2018, 2019, 2020 | 2021, 2022, 2023 |

#### Days Absent

Teachers were asked to indicate the number of days the students in their class have been absent since the beginning of the school year. The number of days absent was categorized into two categories: 0-13 days absent and 14 or more days absent. These categories were based on the current definition of chronic absenteeism, which is defined as missing at least 10% of the school days, or two days a month (35). Given that EDI data were collected between February and April of a given school year, anyone absent for 14 days or more would be considered chronically absent thus far in the school year. An upper limit of 140 days absent was applied to the chronically absent category to align with the number of instructional days that had elapsed at the time of EDI completion.

#### Developmental health

Children’s developmental health outcome was measured using overall developmental vulnerability (i.e. vulnerability in any of the developmental domains of the EDI (36). Teachers rate each child’s abilities and behaviours that are expected of typically developing 5- to 6-year-olds in five general areas of their development using dichotomous (*No*/*Yes*; scored as 0 and 10) or 3-point scales (scored as 0, 5, 10). These five areas are physical health and well-being, social competence, emotional maturity, language and cognitive development, and communication skills and general knowledge. Items on the EDI are then averaged to create a composite score for each developmental domain (ranging from 0 to 10), with higher scores reflecting greater levels of ability in each domain. Using cut-off scores derived from a national baseline collection of EDI data, developmental vulnerability scores (0= *not vulnerable*, 1=*vulnerable*) are then calculated for each domain. Children falling below the 10^th^ percentile cut-off for a given domain are classified as vulnerable in that area. Domain-specific vulnerability scores are then used to calculate an overall developmental vulnerability score: 0= not vulnerable in any domain, 1= vulnerable in at least one domain, which is the variable used in the current study.

#### Child demographics

Child-level demographic information was also collected through the EDI and included: age at time of EDI completion, sex at birth, whether a child has a designation of special needs (SN; yes/no), presence of any functional impairment^2^ (yes/no), and whether the teacher felt the child needed further assessment (yes/no).

#### Neighbourhood-level SES

Information on neighbourhood-level socioeconomic status (SES) was derived using variables from the 2016 Canadian Census and 2015 Taxfiler datasets capturing a range of socioeconomic characteristics. The index used in this study, called the Canadian Neighbourhoods Early Child Development (CanNECD) SES Index (37), is a composite score of 10 variables from these two datasets (Table 2). The SES index was transformed into *Z*-scores (mean of 0 and a standard deviation of 1), with higher SES index representing higher overall neighborhood SES. The CanNECD SES Index was linked to the EDI data using children’s residential postal codes, and was already categorized into quintiles.

**Table 2.**
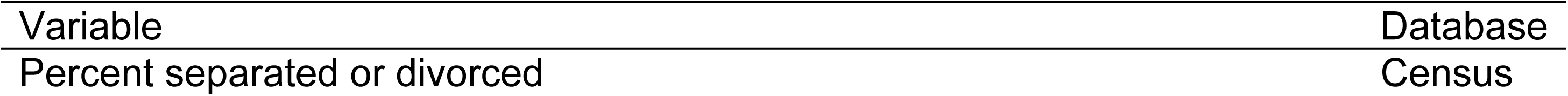
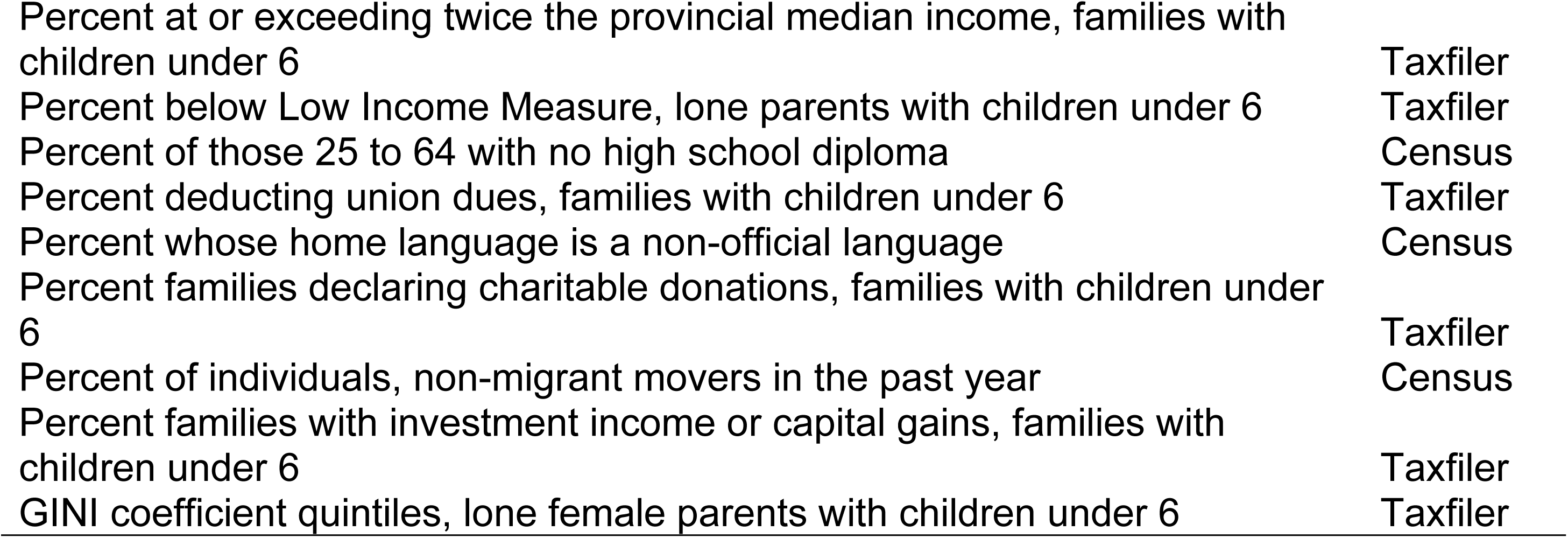
List of the ten socioeconomic variables from the Canadian Census and Taxfiler databases that were included in the CanNECD SES Index.

### Analytic strategy

To start, descriptive statistics including means and proportions were examined for children chronically absent and those who were not, in both pre- and post-COVID-19 onset cohorts. Children’s age, sex, special needs designation, functional impairments, reports of needing further assessment, neighbourhood-level SES, and jurisdiction were compared between children with and without chronic absenteeism, pre- and post-COVID-19 onset using contingency tables. Chi-squared analyses and analyses of variance were conducted to examine differences in sociodemographic characteristics in children not chronically absent and chronically absent children, pre- and post-COVID-19. While statistical significance was considered, because of the size of the study population, emphasis was placed on effect sizes (e.g. Cramer’s V), as they better reflect the magnitude of the differences. Following this, rates of chronic absenteeism between the pre- and post-COVID-19 onset cohorts was compared by conducting an unadjusted binary logistic regression (BLR), followed by adjusted BLRs, controlling for the sociodemographic variables mentioned above, and then including interactions between COVID-19 onset and neighbourhood SES and between COVID-19 onset and jurisdiction. Lastly, to answer the third research question, BLR models were used to examine the association between chronic absenteeism, COVID-19 onset, and developmental vulnerability, first, and then adjusting for the same variables as in the previous BLR models, and last, adjusting for interactions between COVID-19 onset and neighbourhood SES and between COVID-19 onset and jurisdiction.

## Results

### Chronic absenteeism pre- and post-COVID-19

Of a total of 540,005 kindergarten children who were part of the provincial/territorial EDI data collections, 526,850 were considered valid for analysis (97.6%). Of those children, 513,159 (97.4%) were not missing data on any of the variables of interest in this study and were included in the final analytic sample. Of the children included in the analyses, 284,712 (55.5%) were in the pre-COVID-19 cohort and 228,447 (44.5%) were in the post-COVID-19 cohort. Figure 1 shows a flowchart of participants.

**Figure 1.**
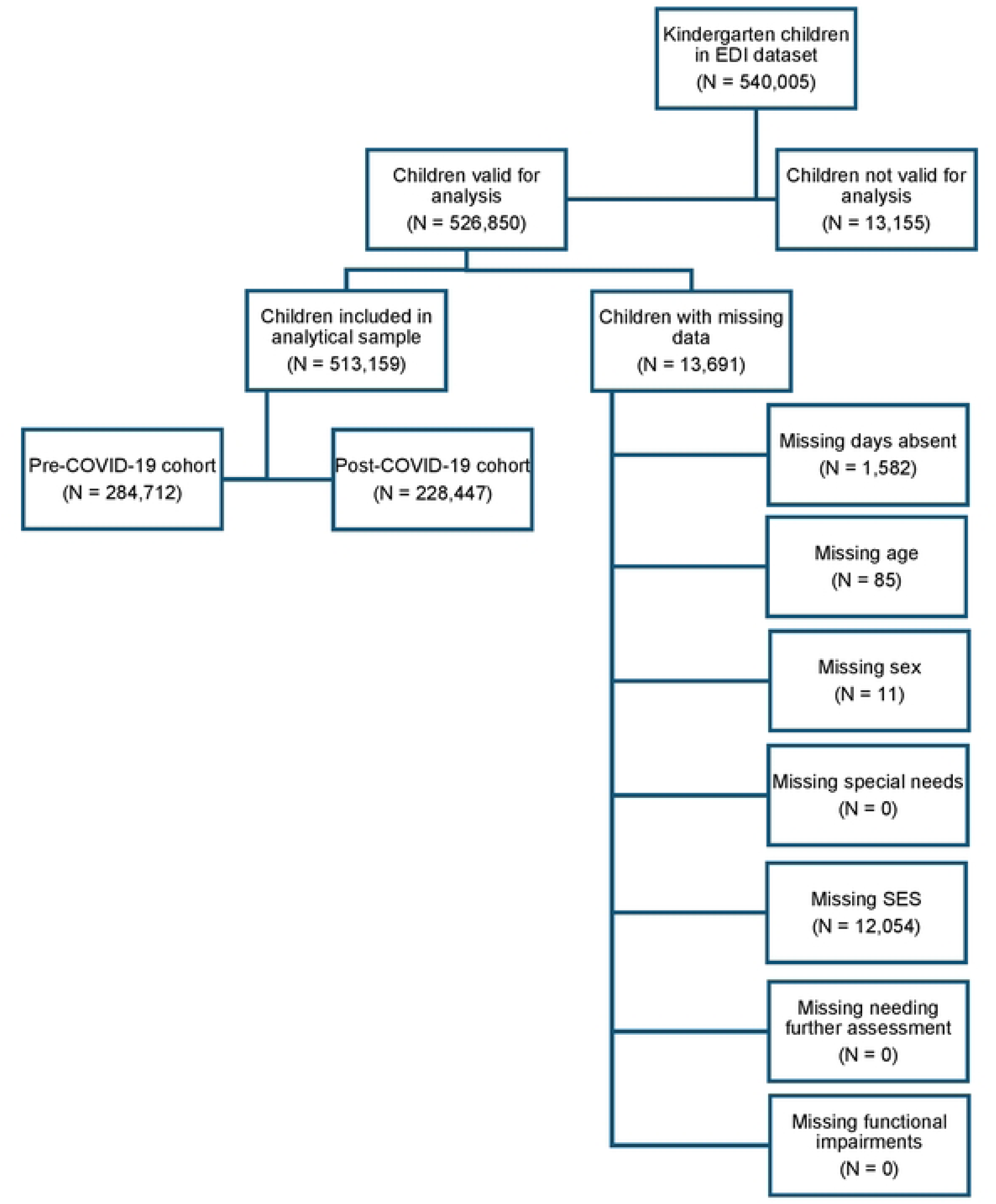
Flowchart of participants

We examined the demographic characteristics of children who were chronically absent and those who were not, pre- and post-COVID-19 (Table 3). As can be seen, most of the demographic characteristics were similar between groups, except for neighbourhood-level SES. There appeared to be a greater proportion of chronically absent students living in higher SES neighbourhoods post-COVID-19 compared to pre-COVID-19.

**Table 3.**
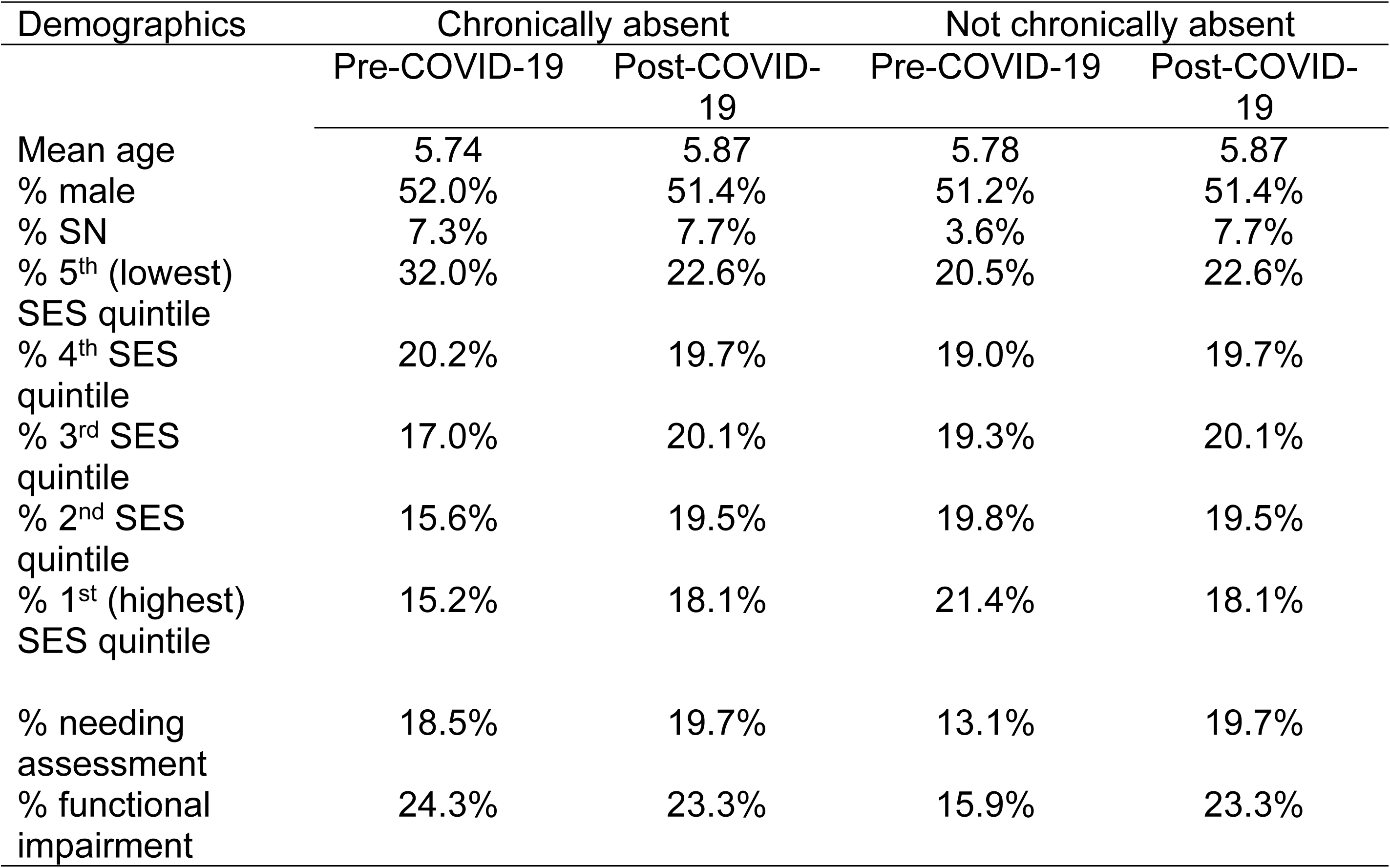
Socio-demographic characteristics of chronically absent kindergarten children in the pre- and post-COVID-19 cohorts, compared to their non-chronically absent peers.

Chi-square tests and analyses of variance were then conducted to examine whether there were differences between the socio-demographic composition of chronically absent children in kindergarten, pre- and post-COVID-19. While statistically significant differences were observed between the two cohorts on all socio-demographic characteristics (Table 4), all effect sizes were small, with the exception of the differences between cohorts for neighbourhood SES and jurisdictions.

**Table 4.** Results of chi-squared tests and analysis of variance examining the socio-demographic composition of chronically absent children, pre- and post-COVID-19 in Canada.

| Demographics | $\chi^2$ | df | *p*-value | Cramer's V |
| --- | --- | --- | --- | --- |
| Sex at birth | 4.22 | 1 | .04 | .005 |
| SN | 6.81 | 1 | .009 | .007 |
| Any FI | 19.46 | 1 | < .001 | .012 |
| Needs assessment | 29.60 | 1 | < .001 | .014 |
| SES | 1720.76 | 4 | < .001 | .109 |
| Jurisdiction | 1939.75 | 7 | < .001 | .116 |
|  | F | df | *p* | Eta-squared |
| Age | 5570.16 | 1 | < .001 | .037 |

$\chi^2$ = chi-square statistic; df = degrees of freedom

Across Canada, rates of chronic absenteeism increased significantly from pre- to post-COVID-19 onset, from 17.7% to 41.3%. Differences in rates of chronic absenteeism were also observed by jurisdiction (Figure 2). The greatest increase was seen in Ontario, while the smallest increase was seen in British Columbia.

**Figure 2.**
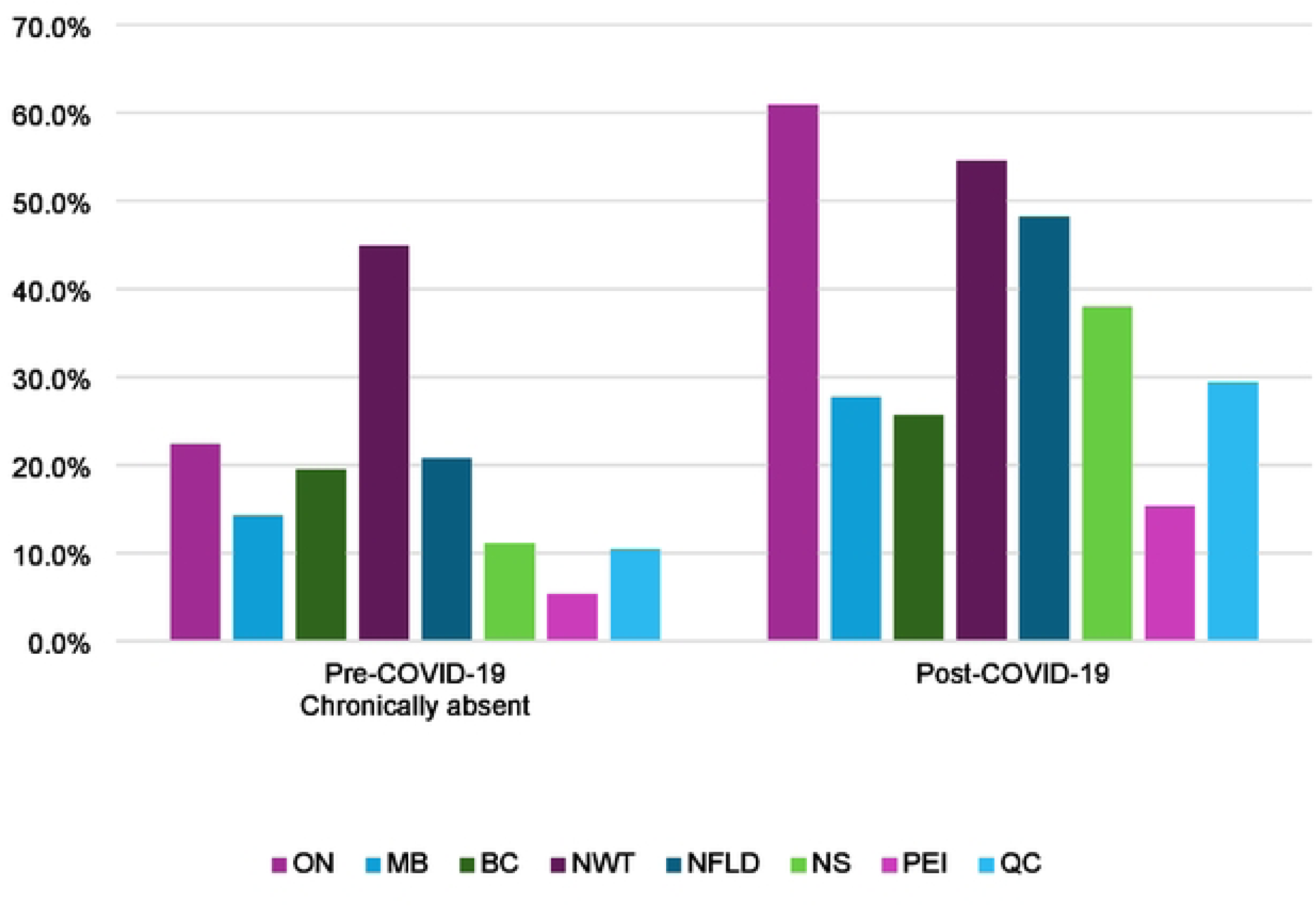
Percentages of kindergarten children chronically absent in each Canadian jurisdiction, pre- and post-COVID-19 onset

Results of an unadjusted BLR revealed that children who attended kindergarten after the onset of the COVID-19 pandemic had 3.26 times greater odds of being chronically absent, compared to their peers who attended kindergarten before the pandemic (Table 5).

**Table 5.** Results of a binary logistic regressions (unadjusted and adjusted) examining the association between pre- and post-COVID-19 cohorts and chronic absenteeism.

| Chronic absenteeism | UOR (95 % CI) *p* | AOR<sup>1</sup> (95% CI) *p* | AOR<sup>2</sup> (95% CI) *p* |
| --- | --- | --- | --- |
| Post COVID-19 | 3.26 (3.21-3.30)<br><.001 | 3.79 (3.74-3.84)<br>< .001 | 3.02 (2.89-3.16)<br>< .001 |

Abbreviations: UOR, unadjusted odds ratio; AOR, adjusted odds ratio; CI, confidence interval.
AOR<sup>1</sup> controlled for sex at birth, age, SN status, presence of functional impairments, needing further assessment, neighbourhood-level SES, and province/territory
AOR<sup>2</sup> controlled for sex at birth, age, SN status, presence of functional impairments, needing further assessment, neighbourhood-level SES, province/territory, and the interactions between COVID-19 onset and SES and between COVID-19 onset and jurisdiction.

After adjusting for the socio-demographic characteristics, including interactions between COVID-19 onset and SES and between COVID-19 onset and jurisdiction, the odds of being chronically absent post-COVID-19 onset changed to 3.02.

### Association between chronic absenteeism and developmental vulnerability, pre-and post-COVID-19

Figure 3 displays the numbers and percentages of children developmentally vulnerable overall, by chronic absenteeism in both cohorts of kindergarten students. A smaller percentage of children chronically absent was vulnerable on one or more developmental domains after the COVID-19 pandemic, compared to their peers attending kindergarten before, whereas rates for non-chronically absent children were fairly similar.

**Figure 3.**
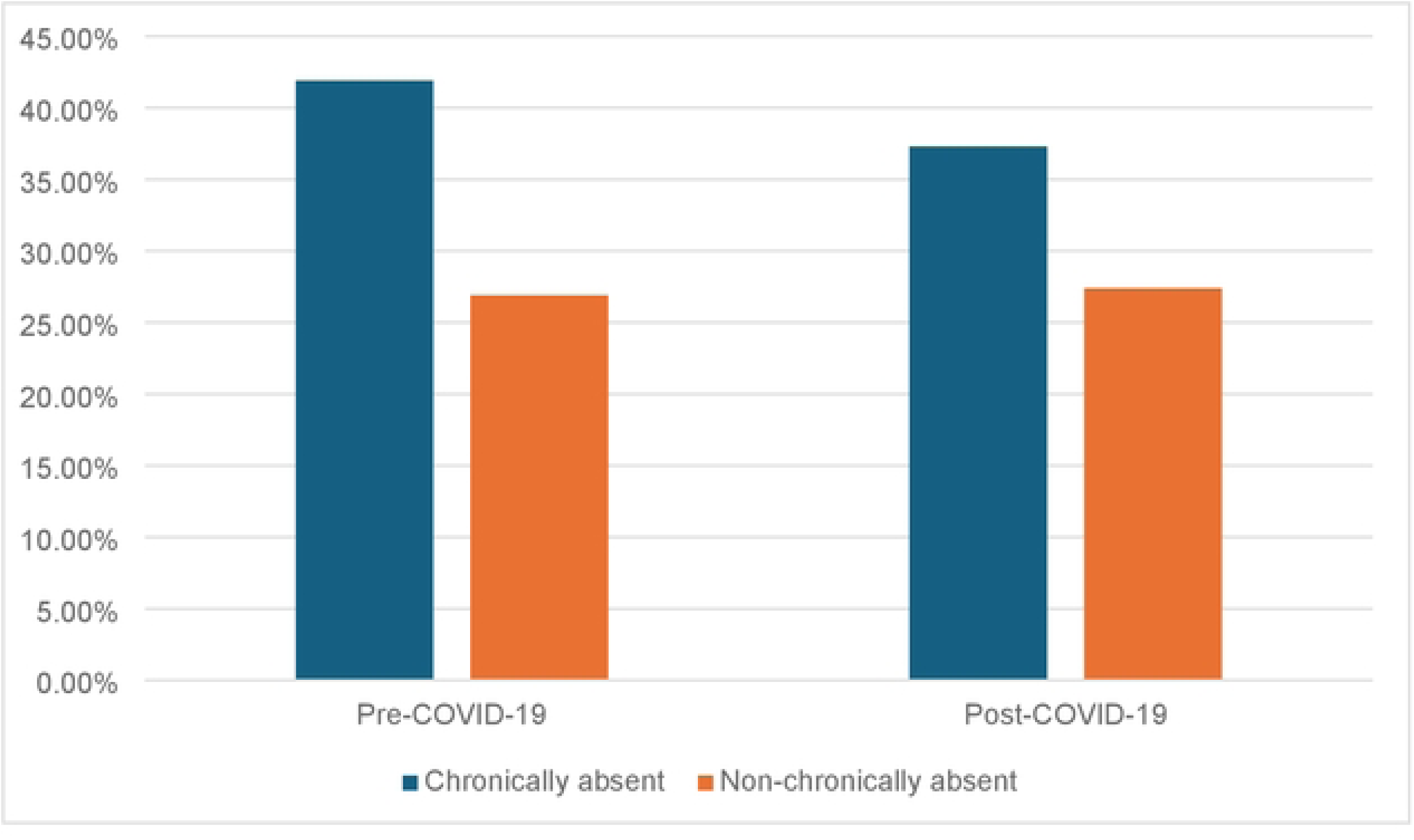
Percentage of children developmentally vulnerable overall by chronic absenteeism, pre- and post-COVID-19

To empirically test the significance of these changes, we conducted an unadjusted BLR to examine the association between chronic absenteeism and developmental vulnerability, pre- and post-COVID-19. We examined the odds of being developmentally vulnerable overall by chronic absenteeism, attending kindergarten post-COVID-19 onset, and by the interaction between these two variables. As can be seen in Table 6, children who were chronically absent from school had 1.96 greater odds of being developmentally vulnerable compared to their peers who were not chronically absent, while children attending kindergarten post-COVID-19 onset had 1.03 greater odds of being developmentally vulnerable compared to their peers who attended school before the pandemic started. Interestingly, the interaction between these two variables resulted in decreased odds of being developmentally vulnerable. Once we controlled for socio-demographic variables, the odds of chronically absent children being vulnerable changed to 1.72, and the odds of being developmentally vulnerable for children attending school post-COVID-19 onset became non-significant. We also observed a reduction in the odds of being developmentally vulnerable post-COVID-19 onset for children who were chronically absent and controlling for other covariates and interactions between COVID-19 onset and SES and between COVID-19 onset and jurisdiction, the reduction in odds of being developmentally vulnerable did not change.

**Table 6.**
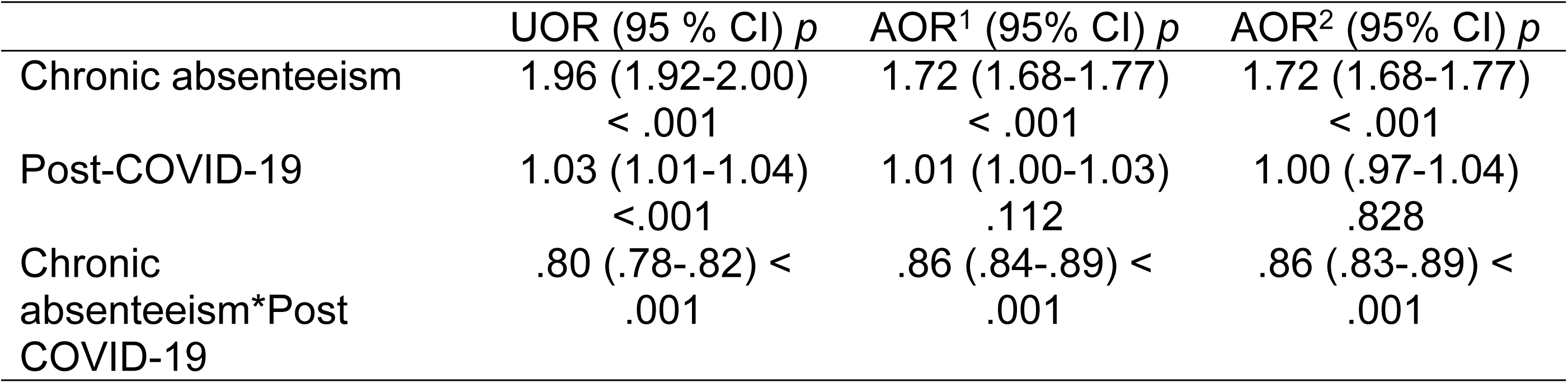
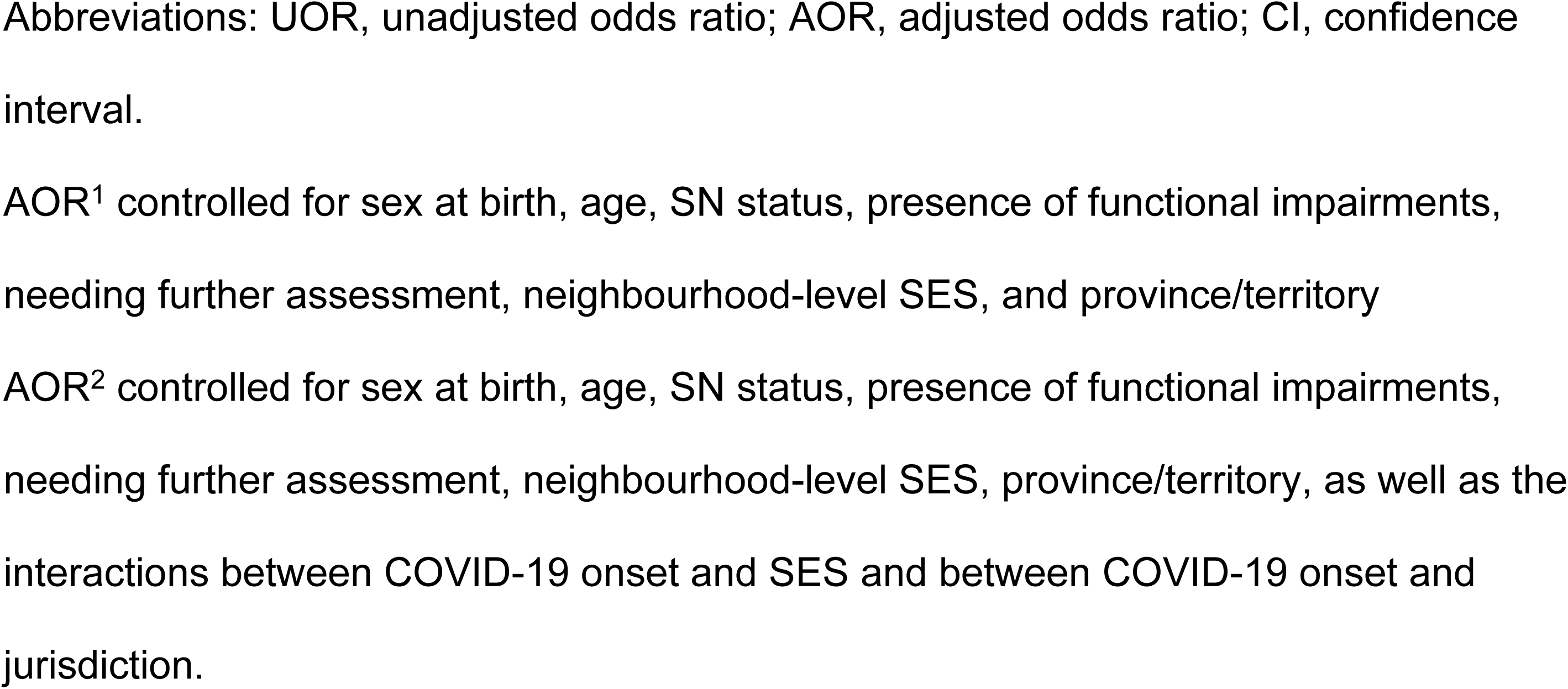
Results of binary logistic regression models examining the association between chronic absenteeism and developmental vulnerability, pre- and post-COVID-19.

## Discussion

In this population-level study of kindergarten children in Canada in a pre- and post-COVID-19 pandemic onset cohorts, we found that children attending school after the onset of the COVID-19 pandemic were three times more likely to be chronically absent compared to their peers attending kindergarten before the pandemic. Changes in rates of chronic absenteeism differed by jurisdiction. Even though its rates increased across the board, chronic absenteeism in the post-COVID-19 period was linked to reduced odds of overall developmental vulnerability – poor school readiness - a pattern that is likely attributable to shifts in the demographic characteristics of chronically absent children. In the post-COVID-19 cohort, a greater percentage of those children lived in higher SES neighbourhoods compared to their chronically absent peers attending school before the onset of the pandemic.

Previous research has reported increases in rates of chronic absenteeism in children in all grades since the onset of the COVID-19 pandemic. For example, rates of chronic absenteeism in publicly-funded schools in the United States increased from approximately 15 percent of students from kindergarten to Grade 12 in 2018-2019 to 28 percent in 2021-22 (25). Even after the pandemic and related-public health measures subsided, rates of chronic absenteeism still remained higher than pre-pandemic levels (38). Our study adds to the evidence on rates of chronic absenteeism increasing after the onset of the COVID-19 pandemic by focusing on the youngest students, who, in the post-pandemic cohort were not even in school during the closures.

Our study demonstrated the highest increase in rates of kindergarten chronic absenteeism was in Ontario, while the smallest increase was in British Columbia. Several factors can potentially explain this difference. Firstly, the jurisdictional differences in rates of chronic absenteeism could be due to the various public health measures put in place. Healthcare and education are predominantly a provincial and territorial responsibility in Canada, and with some exceptions, the majority of pandemic-related public health policy restrictions were introduced by provincial governments (39). For example, Ontario had the greatest number of full and partial school closures in 2020 and 2021 (40), which could have led to challenges with re-engaging in school once in-person instruction resumed. A common assumption is that the increased use of remote instruction in public schools during the 2020–2021 closures contributed to higher chronic absenteeism by interrupting established routines and reducing students’ engagement with schoolwork (25). Increased rates of chronic absenteeism may also be a result of how attendance differed in response to heightened illness and infection risks as students returned to in-person learning. Many schools had strict public health policies when it came to cold and flu symptoms once schools reopened for in-person learning (41), such as dismissing symptomatic students or isolating if symptomatic, which could have resulted in children potentially missing more school compared to before the pandemic when students were more likely to attend school still symptomatic. Furthermore, parental anxiety about illness could also explain an increase in chronic absenteeism since COVID-19. Approximately one-third of parents reported being a little anxious, while 13% and 16% of primary and secondary school parents, respectively, reported being extremely anxious about their children returning to school (42). Hundreds of parents expressed their concerns about their children’s inability to prevent the spread of the disease and their lack of self-consciousness, fearing that children would not respect public health measures, such as wearing their masks while at school (43). Thus, absenteeism may have been influenced by how parents and students reacted to masking policies and other public health measures during the reopening period. Another study examining parents’ willingness to send their children back to school following COVID-19-related school closures found that 31% preferred to keep their children at home, whereas 49% intended to return them to in-person schooling. Decisions about returning to school were influenced by parents’ confidence in the school, the difficulties associated with distance learning, and work-related factors (44), not to mention parents’ concerns about their children’s health and safety (45). Parental attitudes toward school attendance appear to have changed since the pandemic. Since the onset of the pandemic, some parents no longer view daily attendance as essential but instead view it as one of several competing demands in their child’s daily life (46,47).

High rates of school absenteeism are generally seen as an early warning sign for schools, suggesting that children might be struggling either at school or at home (48). Our study showed that, after the onset of the COVID-19 pandemic, chronically absent children had lower odds of being developmentally vulnerable compared to pre-pandemic. Similar findings have also been reported by others (26,49). Chronic school absenteeism tended to be associated with living in poverty or in lower SES areas (6,50). In our study, post-COVID-19, there were a greater proportion of kindergarten children chronically absent from school who were living in higher SES neighbourhoods. This could explain, at least partially, why chronic absenteeism after the onset of the COVID-19 pandemic was associated with decreased odds of developmental vulnerability, as SES is consistently positively associated with children’s developmental health. Several studies have found positive associations between neighbourhood-level SES and children’s developmental outcomes in kindergarten children, where living in higher SES neighbourhoods was associated with better developmental outcomes (37,51,52). It is possible that children from higher SES neighbourhoods missed more school after the onset of the COVID-19 pandemic because their parents had the capability to work from home, making it easier to keep their child(ren) home from school. Work-from-home job postings quadrupled across 20 countries from 2020 to 2023 (53), nevertheless, this increase was not seen across all jobs. Labour market statistics show that the ability to work from home varied, with individuals working in higher income positions and with higher levels of education being more likely to work from home (54).

Another possible explanation for the decreased association between chronic absenteeism and children’s developmental vulnerability post-COVID-19 is that availability and access to online resources has lowered the academic consequences of absenteeism, as students can more readily stay on track with coursework from home (26). In line with this, one research study found that a third (32%) of caretakers were not concerned about their child missing school because, according to them, everything the child needs to know was available online (55). The frequency of children’s participation in academic activities at home during the pandemic, however, varied by parents’ educational attainment. Among parents with at least a bachelor’s degree, 80% reported that their children engaged in structured academic activities at least three times per week, compared to 72% among parents with a college certificate or diploma, 69% among those with a trades certificate or diploma, and 67% among those with a high school diploma or less (56). The reduced strength of association between chronic absenteeism and school readiness in kindergartners after the onset of the COVID-19 pandemic could thus be due to a combined effect of more affluent parents, whose children are more likely to have better developmental outcomes and who have been found to engaged in more structured academic activities at home, and schools providing better resources to absent students for them not to fall back. Future research could address disentangling these possibilities.

### Strengths and limitations

The current study has several strengths. First, we used population-level data from most provinces/territories across Canada, collected over several years, which gave us good insight into chronic absenteeism in kindergarten students across the country before and after the onset of the COVID-19 pandemic. This resulted in absenteeism data for kindergarten children across most of the country, making it the most comprehensive study of the prevalence of chronic absenteeism in young children in Canada to date. Second, the use of a neighbourhood-level SES index initially developed to account for the greatest possible variance in children’s developmental outcomes provided the opportunity to examine the association with SES. Finally, focusing on population-level changes in chronic absenteeism and its association with developmental vulnerability in light of the COVID-19 pandemic provides valuable insight into potential shifts in school absenteeism in cohorts of kindergarten children.

The current study also has limitations. Despite a large sample size and a coverage of two-thirds of Canadian jurisdictions, we were unable to include all jurisdictions of the country, which may decrease the generalizability of our findings. We have little empirical evidence to speculate how post-COVID-19 pandemic outcomes could differ for the regions that did not collect EDI data, and most specifically for territories such as Yukon and Nunavut which have very small populations living in very large, remote regions. We also relied on teacher reports of children’s absenteeism. School boards collect their own attendance data, which may not be collected the same way for all boards/districts, as previously noted (31), potentially leading to differences in the number of days absent reported on the EDI questionnaires. We also did not have information on family-level SES. Finally, to address missing data, we opted to use a complete case analysis. Although multiple imputation is an alternative method for handling missingness, the overall proportion of missing data was small (2.6%), and we therefore proceeded with a complete case approach. We acknowledge, however, that this decision may introduce some degree of bias.

Despite these limitations, this study is an important first step in investigating the prevalence of chronic absenteeism across Canadian kindergarten classes, pre- and post-COVID-19, and its association with children’s developmental vulnerability. Considering our somewhat counter-intuitive findings of decreased association between chronic absenteeism and children’s developmental vulnerability post-COVID-19, future research should examine the associations found in the current study in subsequent cohorts, as well as in primary school among children who did not experience pandemic closures.

## Conclusion

Chronic absenteeism is a well-established early risk factor for school dropout, mental health challenges, financial instability, and academic difficulty (48), often serving as an important early warning sign that students may be experiencing underlying struggles. Although rising rates of chronic absenteeism warrant serious attention, our findings suggest that post-COVID-19 patterns may be more nuanced than in the pre-pandemic period, potentially reflecting shifts in the demographics of students who are chronically absent. The full implications of the pandemic for children’s education and development, however, are still emerging.

A deeper understanding of the factors driving school absences is essential, as are policies aimed at reducing the number of missed days. Because some degree of legitimate absenteeism is inevitable, strategies that mitigate the learning consequences of missed instruction may also be valuable. This area has, however, received limited research and policy focus to date. Supporting schools as they navigate the many challenges intensified or introduced by the pandemic, including chronic absenteeism, will require thoughtful and sustained approaches to school attendance. Such efforts will be critical for promoting children’s academic and socioemotional well-being.

## Data Availability

Data is available upon written request to the Offord Centre for Child Studies

## Acknowledgments

We would like to extend our gratitude to our provincial and territorial partners for their hard work and commitment to the EDI data collections, as well as to the teachers across Canada who committed their time and energy in completing the EDI questionnaires and who have made this work possible. We would also like to thank Paul Lefebvre for his assistance with the analysis and interpretation of the results of the study.

## Footnotes

1 Over 90 % of Canadian children attend publicly-funded schools (32).

2 Teachers are asked to indicate whether they feel a given student in their classroom has a problem that influences their ability to participate in the classroom. If they say yes, they can choose from a list of 11 different impairments, ranging from visual and hearing impairments, to emotional and behavioural problems.

## References

1. Duncan GJ, Dowsett CJ, Claessens A, Magnuson K, Huston AC, Klebanov P, et al. School readiness and later achievement. Dev Psychol. 2007 Nov;43(6):1428–46.

2. Entwistle DR, Alexander KL, Olson LS. Keep the Faucet Flowing. Educ Healthc Public Serv [Internet]. 2001 [cited 2025 Jun 16];25(3). Available from: https://www.aft.org/ae/fall2001/entwisle_alexander_olson

3. Ansari A, Purtell KM. Absenteeism in Head Start and Children’s Academic Learning. Child Dev. 2018 Jul;89(4):1088–98.

4. Gershenson S, Jacknowitz A, Brannegan A. Are Student Absences Worth the Worry in U.S. Primary Schools? Educ Finance Policy. 2017 Apr 1;12(2):137–65.

5. Gottfried MA. Chronic Absenteeism and Its Effects on Students’ Academic and Socioemotional Outcomes. J Educ Stud Placed Risk JESPAR. 2014 Apr 3;19(2):53–75.

6. Morrissey TW, Hutchison L, Winsler A. Family income, school attendance, and academic achievement in elementary school. Dev Psychol. 2014 Mar;50(3):741–53.

7. Smerillo NE, Reynolds AJ, Temple JA, Ou SR. Chronic absence, eighth-grade achievement, and high school attainment in the Chicago Longitudinal Study. J Sch Psychol. 2018 Apr;67:163–78.

8. Davies S, Janus M, Duku E, Gaskin A. Using the Early Development Instrument to examine cognitive and non-cognitive school readiness and elementary student achievement. Early Child Res Q. 2016;35:63–75.

9. Attendance Works, Everyone Graduates Center. Preventing missing opportunities: Taking collective action to confront chronic absence [Internet]. 2016 p. 36. Available from: https://www.attendanceworks.org/wp-content/uploads/2017/04/Preventing-Missed-Opportunity-Full_FINAL9_8_16_2.pdf

10. Chang H, Romero M. Present, Engaged, and Accounted For: The Critical Importance of Addressing Chronic Absence in the Early Grades – NCCP [Internet]. New York, NY, US: National Centre for Children in Poverty, Mailman School of Public Health, Columbia University; 2008 [cited 2025 Jun 18] p. 31. Available from: https://www.nccp.org/publication/present-engaged-and-accounted-for-the-critical-importance-of-addressing-chronic-absence-in-the-early-grades/

11. Heckman JJ. Schools, Skills, and Synapses. Econ Inq. 2008 Jun;46(3):289.

12. Eaton DK, Brener N, Kann LK. Associations of health risk behaviors with school absenteeism. Does having permission for the absence make a difference? J Sch Health. 2008 Apr;78(4):223–9.

13. Hickman GP, Bartholomew M, Mathwig J, Heinrich RS. Differential Developmental Pathways of High School Dropouts and Graduates. J Educ Res. 2008;102(1):3–14.

14. Connolly F, Olson LS. Early Elementary Performance and Attendance in Baltimore City Schools’ Pre-Kindergarten and Kindergarten [Internet]. Baltimore Education Research Consortium; 2012 Mar [cited 2025 Nov 14]. Available from: https://eric.ed.gov/?id=ED535768

15. Ehrlich SB, Gwynne JA, Pareja AS, Allensworth E, Moore PT, Jagesic S, et al. Preschool Attendance in Chicago Public Schools [Internet]. Chicago, IL: UChicago Consortium on School Research; 2014 [cited 2025 Nov 14]. Available from: https://consortium.uchicago.edu/publications/preschool-attendance-chicago-public-schools-relationships-learning-outcomes-and-reaso-0

16. Allen CW, Diamond-Myrsten S, Rollins LK. School Absenteeism in Children and Adolescents. Am Fam Physician. 2018 Dec 15;98(12):738–44.

17. Balfanz R, Byrnes V. The Importance of Being in School: A Report on Absenteeism in the Nation’s Public Schools [Internet]. 2012 p. 44. Available from: chrome-extension://efaidnbmnnnibpcajpcglclefindmkaj/https://new.every1graduates.org/wp-content/uploads/2012/05/FINALChronicAbsenteeismReport_May16.pdf

18. Education Commission of the States. Chronic Early Absence. Prog Eductaion Reform. 2010;11(1):6.

19. Chang HN, Davis R. Mapping the Early Attendance Gap [Internet]. Attendance Works; 2015 p. 36. Available from: https://www.attendanceworks.org/wp-content/uploads/2017/05/Mapping-the-Early-Attendance-Gap_Final-4.pdf

20. Gottfried MA. Quantifying the Consequences of Missing School: Linking School Nurses to Student Absences to Standardized Achievement. Teach Coll Rec. 2013 Jun 1;115(6):1–30.

21. Gottfried MA. Can Neighbor Attributes Predict School Absences? Urban Educ. 2014 Mar 1;49(2):216–50.

22. Gottfried MA, Gee KA. Identifying the Determinants of Chronic Absenteeism: A Bioecological Systems Approach. Teach Coll Rec. 2017 Jul 1;119(7):1–34.

23. Ready DD. Socioeconomic Disadvantage, School Attendance, and Early Cognitive Development: The Differential Effects of School Exposure. Sociol Educ. 2010 Oct 1;83(4):271–86.

24. Van Eck K, Johnson SR, Bettencourt A, Johnson SL. How school climate relates to chronic absence: A multi–level latent profile analysis. J Sch Psychol. 2017 Apr 1;61:89–102.

25. Dee TS. Higher chronic absenteeism threatens academic recovery from the COVID-19 pandemic. Proc Natl Acad Sci. 2024 Jan 16;121(3):e2312249121.

26. Swiderski T, Fuller SC, Bastian KC. The Relationship Between Student Attendance and Achievement, Pre- and Post-COVID. AERA Open. 2025 Sep 1;11:23328584251371041.

27. Malkus N. Lingering Absence in Public Schools: Tracking Post-Pandemic Chronic Absenteeism into 2024 [Internet]. American Enterprise Institute - AEI. 2025 [cited 2025 Dec 8]. Available from: https://www.aei.org/research-products/report/lingering-absence-in-public-schools-tracking-post-pandemic-chronic-absenteeism-into-2024/

28. Gagné M, Janus M, Milbrath C, Gadermann A, Guhn M. Early emotional and communication functioning predicting the academic trajectories of refugee children in Canada. Educ Psychol. 2018 Sep 14;38(8):1050–67.

29. Guhn M, Gadermann AM, Almas A, Schonert-Reichl KA, Hertzman C. Associations of teacher-rated social, emotional, and cognitive development in kindergarten to self-reported wellbeing, peer relations, and academic test scores in middle childhood. Early Child Res Q. 2016 Apr 1;35:76–84.

30. Carman T, Wesley A. Kids missing more school since pandemic, CBC analysis finds. CBC News [Internet]. 2024 Mar 27 [cited 2025 May 30]; Available from: https://www.cbc.ca/news/canada/school-absence-data-1.7156254

31. Carman T. Why measuring school absences in Canada is so hard. CBC News [Internet]. 2025 Nov 22 [cited 2025 Dec 4]; Available from: https://www.cbc.ca/news/canada/school-absences-data-9.6988718

32. Statistics Canada. Elementary–Secondary Education Survey, 2020/2021 [Internet]. 2022 [cited 2025 Aug 27]. Available from: https://www150.statcan.gc.ca/n1/daily-quotidien/221013/dq221013a-eng.htm

33. Janus M, Offord DR. Development and psychometric properties of the Early Development Instrument (EDI): A measure of children’s school readiness. Can J Behav Sci Rev Can Sci Comport. 2007;39(1):1–22.

34. Janus M, Gaskin A, Reid-Westoby C. School Readiness. In: Encyclopedia of Quality of Life and Well-Being Research [Internet]. Springer, Cham; 2022 [cited 2025 Dec 18]. p. 1–6. Available from: https://link.springer.com/rwe/10.1007/978-3-319-69909-7_2600-2

35. U.S. Department of Education. Chronic Absenteeism [Internet]. 2025 [cited 2025 Jun 19]. Available from: http://www.ed.gov/teaching-and-administration/supporting-students/chronic-absenteeism

36. Janus M, Duku E. The school entry gap: Socioeconomic, family, and health factors associated with children’s school readiness to learn. Early Educ Dev. 2007 Oct 11;18(3):375–403.

37. Forer B, Minh A, Enns J, Webb S, Duku E, Brownell M, et al. A Canadian neighbourhood index for socioeconomic status associated with early child development. Child Indic Res. 2020 Aug 1;13(4):1133–54.

38. Malkus N. Long COVID for Public Schools: Chronic Absenteeism before and after the Pandemic [Internet]. American Enterprise Institute; 2024 Jan [cited 2025 Sep 10]. Available from: https://eric.ed.gov/?id=ED674001

39. Cameron-Blake E, Breton C, Sim P, Tatlow H, Wood A, Smith J, et al. Variation in the Canadian provincial and territorial responses to COVID-19. Blavatnik Sch Gov Work Pap [Internet]. 2021; Available from: www.bsg.ox.ac.uk/covidtracker

40. Akanteva A, Dick DW, Amiraslani S, Heffernan JM. Canadian Covid-19 pandemic public health mitigation measures at the province level. Sci Data. 2023 Dec 8;10(1):882.

41. Public Health Ontario. Environmental Scan: School-based publich health measures in select jurisdictions and guidance from public health organizations [Internet]. 2022. Available from: https://www.publichealthontario.ca/-/media/Documents/nCoV/phm/2022/08/school-public-health-measures-pho-guidance.pdf?rev=a4b8227db0c6426dacd74a92d4b0f2e2&sc_lang=en

42. Amin-Chowdhury Z, Bertran M, Kall M, Ireland G, Aiano F, Powell A, et al. Parents’ and teachers’ attitudes to and experiences of the implementation of COVID-19 preventive measures in primary and secondary schools following reopening of schools in autumn 2020: a descriptive cross-sectional survey. 2022 Sep 1 [cited 2025 Sep 26]; Available from: https://bmjopen.bmj.com/content/12/9/e052171.abstract

43. Zhan Z, Li Y, Yuan X, Chen Q. To Be or Not to Be: Parents’ Willingness to Send Their Children Back to School After the COVID-19 Outbreak. Asia-Pac Educ Res. 2022 Oct 1;31(5):589–600.

44. Hageman JR. Can Students Safely Return to School in the Age of COVID-19? Pediatr Ann. 2020 Sep;49(9):e363–4.

45. Limbers CA. Factors Associated with Caregiver Preferences for Children’s Return to School during the COVID-19 Pandemic. J Sch Health. 2021;91(1):3–8.

46. Lester KJ, Michelson D. Perfect storm: emotionally based school avoidance in the post-COVID-19 pandemic context. BMJ Ment Health [Internet]. 2024 Apr 5 [cited 2025 Jun 4];27(1). Available from: https://mentalhealth.bmj.com/content/27/1/e300944

47. Weale S. Parents in England no longer see daily school attendance as vital, report finds. The Guardian [Internet]. 2023 Sep 20 [cited 2026 Jan 7]; Available from: https://www.theguardian.com/education/2023/sep/21/parents-in-england-no-longer-see-daily-school-attendance-as-vital-report-finds

48. Kearney CA, Dupont R, Fensken M, Gonzálvez C. School attendance problems and absenteeism as early warning signals: review and implications for health-based protocols and school-based practices. Front Educ [Internet]. 2023 Aug 30 [cited 2025 May 30];8. Available from: https://www.frontiersin.org/journals/education/articles/10.3389/feduc.2023.1253595/full

49. Fuller SC, Swiderski T, Mikkelsen C, Bastian KC. In School, Engaged, on Track? The Effect of the Pandemic on Student Attendance, Course Grades, and Grade Retention in North Carolina. Educ Res. 2025 Mar 1;54(2):78–90.

50. Gottfried MA. Can Neighbor Attributes Predict School Absences? Urban Educ. 2014 Mar 1;49(2):216–50.

51. Halfon N, Aguilar E, Stanley L, Hotez E, Block E, Janus M. Measuring Equity From The Start: Disparities In The Health Development Of US Kindergartners. Health Aff (Millwood). 2020 Oct 1;39(10):1702–9.

52. Janus M, Reid-Westoby C, Raiter N, Forer B, Guhn M. Population-Level Data on Child Development at School Entry Reflecting Social Determinants of Health: A Narrative Review of Studies Using the Early Development Instrument. Int J Environ Res Public Health. 2021 Jan;18(7):3397.

53. Adrjan P, Ciminelli G, Judes A, Koelle M, Schwellnus C, Sinclair TM. Working from home after COVID-19: Evidence from job postings in 20 countries. Labour Econ. 2025 Oct 1;96:102751.

54. Government of Canada. Running the economy remotely: Potential for working from home during and after COVID-19 [Internet]. 2020 [cited 2025 Sep 22]. Available from: https://www150.statcan.gc.ca/n1/pub/45-28-0001/2020001/article/00026-eng.htm

55. Saavedra A, Polifkoff M, Silver D. Parents are not fully aware of, or concerned about, their children’s school attendance [Internet]. Brookings. 2024 [cited 2025 Sep 26]. Available from: https://www.brookings.edu/articles/parents-are-not-fully-aware-of-or-concerned-about-their-childrens-school-attendance/

56. Government of Canada. School closures and COVID-19: Interactive Tool [Internet]. 2021 [cited 2025 Nov 18]. Available from: https://www150.statcan.gc.ca/n1/pub/71-607-x/71-607-x2021009-eng.htm

